# What is expected of people who lead meetings where the goal is to reach consensus? A scoping review with implications for improving the quality of health research grant peer review and clinical guideline development

**DOI:** 10.1101/2025.02.03.25320968

**Authors:** Mona Ghannad, Anna Catharina V. Armond, Jeremy Y Ng, Hassan Khan, Dean Giustini, Anne Lasinsky, Joanie Sims-Gould, Paul Blazey, Nadia Martino, Sammy Nag, Adrian Mota, David Moher, Karim M Khan, Clare L Ardern

## Abstract

**Background:** The specific roles and responsibilities expected of leaders of consensus-based decision committees, such as grant peer review panels and guideline development panels, are not well-defined, which makes it difficult to train people to lead well. We aimed to explore, describe and define the roles, responsibilities, and leadership characteristics of leaders of meetings where the goal was to reach a consensus decision.

**Methods:** We conducted a scoping review with thematic synthesis, guided by the Joanna Briggs Institute Scoping Review Methodology, and Arksey & O’Malley’s framework for scoping reviews as refined by Levac et al. We searched five bibliographic databases from January 2002-2023 in English: Medline (Ovid), Embase (Ovid), CINAHL (EBSCO) and PsycInfo (EBSCO); Proquest Digital Dissertations and ABI-Inform. We searched grey literature in the fields of health science, biomedicine, education, psychology, management, law, ethics and policy. Abstracts and full-text articles were screened in duplicate to identify eligible studies; data were extracted regarding the roles, responsibilities and characteristics of consensus decision committee leaders. Themes were constructed using reflexive thematic analysis.

**Results:** From 6732 electronic database records and 126 grey literature records, we included 24 articles and 16 websites. There were 166 unique statements extracted related to roles and responsibilities. We constructed 4 themes to describe the roles for leaders of consensus-based decision meetings: (1) *organizer and/or resource manager*, (2) *facilitator*, (3) *adjudicator* and, (4) *administrator*.

**Conclusion:** Leaders of consensus committees assumed the roles of organiser and/or resource manager, facilitator, adjudicator and administrator. Better clarification of and training for the expected roles and responsibilities of leading consensus decisions are needed. Establishing the roles and responsibilities can inform a systematic process for evaluating the performance of leaders of consensus decision committees.

## Introduction

Leaders of consensus decision meetings (e.g., chairs of grant peer review or clinical guideline panels) can wield great influence over those they lead. In an analysis of guideline panels’ decision-making, the chair and co-chair combined accounted for two-thirds of the conversations.^1^ Developed in the 1980s, the RAND-UCLA approach to reaching consensus about the appropriateness of performing a medical procedure is widely used, including in fields outside medical decision-making. RAND-UCLA’s guidance underscores the crucial role of a leader (moderator) figure in driving the consensus process and influencing decisions.^2^

Our research group is interested in the processes of grant peer review—how it is conducted and evaluated—and in the science of how consensus is reached among guideline panel members.^3–7^ Studying these processes matters because they are used by most national research funders to select which grant applications to fund,^8^ ^8^ ^9^ and by guideline panels to make recommendations that influence important healthcare decisions.^10^ Yet the integrity of peer review is increasingly questioned and historically prone to bias, and poor quality and reliability of peer review is a problem.^8^ ^9^ ^11^ ^12^ The recent ACCORD Guideline emerged from a need to improve reporting standards and rigour in consensus methods, including for developing clinical guidelines.^10^ ^13^

Sometimes the loudest voices prevail and the opinions and perspectives of the committee’s leaders (e.g., chairs) are valued more than other committee’s members.^14^ We have found it is unusual for leaders of consensus committees to receive structured training.^6^ Evaluating the performance of groups that are tasked with reaching consensus decisions, including grant peer review panels, is crucial to improving the quality of consensus. Evaluating and reporting on performance can provide transparency regarding how consensus committees function, and help identify problem areas for which training modules could be developed.

To help consensus decision committees succeed, consensus leaders need to know what is expected of them (i.e. roles and responsibilities), and to receive specific training aimed at supporting leaders to fulfill the roles and responsibilities. We suspect that consensus leaders may not receive sufficient/adequate training, and we contend that defining the roles and responsibilities would guide quality improvement efforts.^15^ We see at least 3 important questions that are worthy of study as a first step towards creating structures and training to support consensus leaders to deliver a quality, transparent and equitable process: 1) how are the roles and responsibilities of consensus leaders defined and described?, 2) what specific training do consensus leaders need to excel in their roles? and, 3) how should organisations best evaluate the performance of consensus leaders?

The aims of our scoping review were constructed to deliver key information to underpin training materials for consensus decision committee leaders. We aimed to:

1. Describe and define the *roles* of leaders of meetings where the goal was to reach a consensus-based decision.
2. Describe and define the *responsibilities* of leaders of meetings where the goal was to reach a consensus-based decision
3. Explore the leadership characteristics of consensus decision committee leaders

## Methods

We conducted a scoping review with qualitative synthesis using reflexive thematic analysis,^16^ guided by the Joanna Briggs Institute Scoping Review Methodology Group^17^ approach, and Arksey & O’Malley’s framework for scoping reviews^18^ as refined by Levac et al.^19^ The review protocol was prospectively registered on the Open Science Framework.^20^ We briefly outline our review methods here, referring to the review protocol; there were no deviations from the protocol as prospectively registered. Our scoping review is reported according to the Preferred Reporting Items for Systematic reviews and Meta-Analysis – Scoping Review extension guidelines.^21^

### Searching

Evidence regarding the role of committee chairs and consensus is disseminated in academic journals and monographs.^22^ ^23^ An experienced medical librarian (DG) guided a comprehensive search strategy of bibliographic databases. We used a combination of subject headings, free-text and keyword phrases. We performed scoping searches to estimate the literature’s size and tested the sensitivity and specificity of the search strategies in monographs (using Proquest Digital Dissertations and ABI-Inform), grey literature, and in 5 bibliographic databases: Medline (Ovid), Embase (Ovid), CINAHL (EBSCO) and PsycInfo (EBSCO). A gold standard search was developed for Medline and adapted to the other databases (Appendix 1).

To supplement the bibliographic database search, we searched the Web of Science Core Collection and Scopus citation indices for relevant papers using the major concepts, and used forward and backward referencing of relevant systematic reviews to identify any further relevant literature. Our pragmatic approach to balancing the sensitivity of the search with the capacity of the research team meant that we limited our searches to studies published from January 2002 in English. The searches were first run on 6 January 2023, and updated on 29 October 2024.

We searched the grey literature in March 2023 (Appendix 2). We performed keyword searches using the native search functions on the homepages of the world’s 50 largest biomedical research funders (as determined by amount of annual research funding available, and listed at https://www.healthresearchfunders.org/), including government and philanthropic organisations. We searched the websites of other academic bodies (e.g., learned societies) and relevant web sources (e.g., management research organizations) that we expected might have published relevant research characterising the roles and responsibilities of organisational chairs and management leaders. The search terms (keywords) were: “leaders”, “chair”, “scientific officer”, “roles”, “responsibilities”.

### Eligibility criteria

Our search focused on identifying studies that reported on leadership roles within grant peer review committees, journal peer review committees, and decision committees across small or large for-profit and not-for-profit organisations. We sought data from empirical studies from the disciplines of: health science, biomedicine, education, psychology, management research, law, ethics, and policy. ^24^

### Inclusion criteria

We included articles, interviews, reports, or other digital documents, including statements, guidance, mandates, or policies, that reported on 1) roles, 2) responsibilities, 3) criteria for evaluating the performance of consensus committee leaders, or 4) characteristics of effective leadership of consensus meetings.

Consensus decisions are reached when a group of people work towards agreeing on a course of action prior to finalising a decision.^25^ We included records that described consensus decision committees, where the committees followed a similar process to grant peer review: comprising approximately 10-30 members (this number is an approximation and was not used as a selection criterion as journal peer review committees may have fewer members), with a designated primary and secondary leadership role (e.g., chair and vice chair/scientific officer) managing and facilitating the meeting, where the goal was to reach a decision within a group setting. We included social network analysis (SNA), as these records offered value in understanding leadership roles in consensus decision-making committees.

### Exclusion criteria

We excluded literature that was not published in English; editorials, commentaries or opinion papers, case reports (n-of-1 studies), conference proceedings, and narrative reviews. We excluded legislative debates or legal/court of law arguments, reports of advisory groups (where the decision-making focus was on allocating tasks), or meetings between adversary groups (e.g., political campaigns). We excluded literature where executive committees of only high-ranking members (e.g., chief officers, governors, directors or trustees), or where all members held equal positions of authority and there was no designated committee leader. We also excluded teaching materials and courses offered in leadership and management research, as they focused on how to lead, not on the decision process.

### Study selection

Search results were downloaded and saved in EndNote 20.4 *(Clarivate Analytics, PA, USA)*, then deduplicated ^26^ and uploaded to Covidence (Veritas Health Innovation, Melbourne, Australia) for screening.

We piloted the selection criteria using a random sample of 50 records (titles and abstracts) of studies obtained from the search by two independent reviewers. We aimed for agreement of at least 90% during the pilot screening phase. After piloting was complete, pairs of independent reviewers screened the titles and abstracts to identify potentially eligible studies. For full text screening, pairs of reviewers independently screened all potentially relevant full-text records for eligibility. Disagreements were resolved by consensus or by a third member of the research team, as required.

### Data extraction

Data were extracted by one reviewer and verified by a second reviewer using a custom Microsoft Excel template.^20^ Conflicts were resolved by consensus or when necessary, by a third researcher. We extracted study characteristics, including publication year and data source, and potential performance indicators related to the roles and responsibilities of consensus leaders, and key attributes of the leaders. Qualitative data included statements or descriptions related to roles and/or responsibilities (before, after, or during meetings), criteria for assessing performance, behaviours of consensus leaders, accountability (e.g., the person and/or department the consensus leader reported to), tenure (e.g., time served), time commitment, compensation, conflict of interest, and any other relevant outcomes.

### Data analysis: thematic synthesis

Reflexive thematic analysis, with an inductive approach that was informed by interpretivist and constructivist paradigms, was used to analyse and synthesise the qualitative data.^27–30^ A reflexive approach to thematic synthesis was appropriate because we aimed to identify patterns and themes in the data regarding roles, expectations and behaviours of consensus decision committee leaders.

The analysis phases were: reading and re-reading the extracted text (familiarising), generating initial codes, generating themes, reviewing potential themes, defining and naming themes, and report writing.^27–29^ ^31^ One researcher took a dynamic and recursive approach to generating codes and themes from the data. Reflecting a subjective approach, while constructing the codes and themes, the researcher collaborated with two other research team members in a reflexive manner for sense-checking of ideas, and to explore multiple interpretations of the data. The collaborative and reflexive approach aimed to reach a richer interpretation of the data, not to reach consensus on the meaning.^28^ ^29^ Extracts from the data items underlying each theme (data extracts) were constructed to describe and define the roles and responsibilities of consensus decision committee leaders. We present the data extracts descriptively.^16^ ^28–30^

## Results

The bibliographic database search retrieved 9547 records; from the grey literature, we identified 106 potentially relevant websites and added 20 highly-relevant citations from pilot searches. After removing duplicates, there were 6732 records included for title and abstract screening, and we excluded 6527 records that did not meet the eligibility criteria (Figure 1).

**Figure 1.**
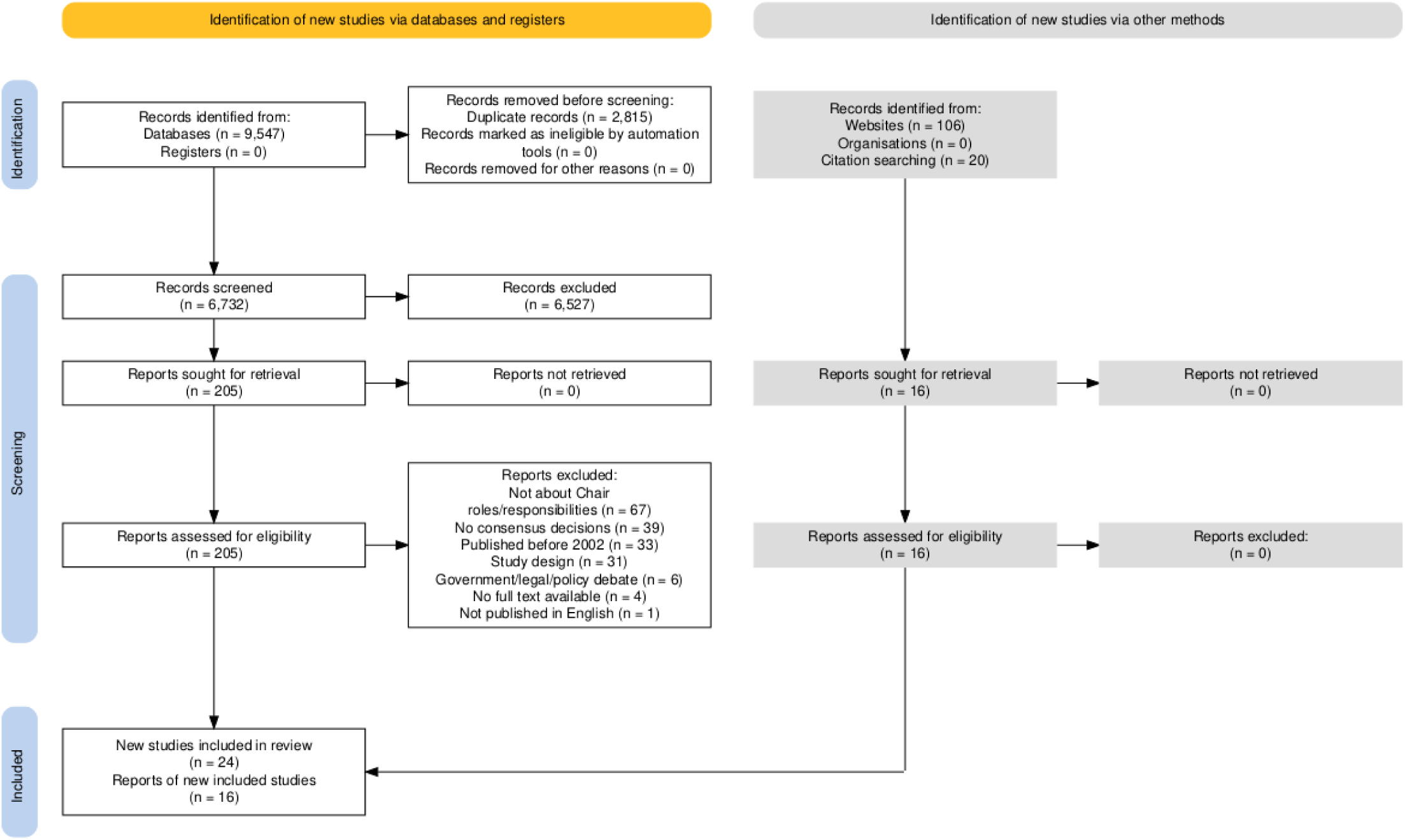
Flow chart of search records.

We reviewed 205 records in full text, and included 24 records for review (Figure 1).^14^ ^32–54^ We excluded 181 records for the following reasons: roles/responsibilities of meeting leaders not reported (n = 67), consensus decisions not studied (n = 39), published prior to 2002 (n = 33), ineligible study design (n = 31), government/legal/policy debates or meetings between adversary groups (n = 6), no full text available (n = 4), published in a language other than English (n = 1). The grey literature search identified 106 potential records, from which we included 16 records (Figure 1).^55–70^

### Roles and responsibilities of consensus decision committee leaders

We extracted 166 statements related to roles and responsibilities (68 statements describing roles, 60 statements describing responsibilities, and 38 statements describing general roles and responsibilities) from the 40 included records.

We constructed 4 themes to reflect the roles and associated responsibilities of consensus decision committee leaders: 1) organiser, 2) facilitator, 3) adjudicator, and 4) administrator (Table 1).

**Table 1.** Roles and responsibilities of consensus decision committee leaders.

| **Role** | **Description** | **Responsibilities** |
| --- | --- | --- |
| Organiser | The person responsible for adhering to the logistic and technical aspects of the meetings. | • Identify/select committee members<br>• Direct what is required of committee members (articulate rules/expectations and agenda)<br>• Identify content experts as necessary<br>• Manage the meeting timing, flow and questions<br>• Maintain a confidential process<br>• Actively manage conflicts of interest |
| Facilitator | The person responsible for managing the meeting interactions. | • Ensure rules and procedures are adhered to<br>• Ensure all committee members have the necessary background information to participate in discussions<br>• Balance adherence to the meeting agenda<br>• Provide opportunities for all members to participate<br>• Recognise and manage interpersonal conflict |
| Adjudicator | The person responsible for ensuring balance and fairness in committee discussions. | • Ensure discussion of scientific and technical merit of information considered by the committee<br>• Synthesise information<br>• Discuss and mediate discrepant opinions/ratings<br>• Manage conflict<br>• Recognise and limit the influence of potential sources of bias<br>• Direct deliberations towards a decision (consensus) and bring discussions to a close |
| Administrator | The person responsible for governance and record keeping. | • Record detailed meeting notes<br>• Prepare summaries of the discussion<br>• Flag issues related to ethics, budgets etc. for discussion<br>• Document and help manage conflicts of interest |

Each role had specific responsibilities that occurred at every stage of the consensus decision committee meeting (i.e., before, during, and after). The data extracts presented below are intended to illustrate each role and associated responsibilities.

### The organiser

The *organiser* co-ordinated the logistics and technical aspects of consensus decision meetings;^44^ ^56^ keeping the consensus meeting on track (planning for, managing and conducting the meeting) and managing the resources needed for a productive meeting.^38^ ^41^ ^46^ ^55^ ^56^ ^59^ ^63^ Prior to a consensus decision meeting, the *organiser* prepared and distributed materials (including pre-reading), helped to select and recruit consensus decision committee members, identified appropriate content experts who would provide specific expertise not held within the consensus decision committee and directed consensus decision committee members to follow meeting guidelines.^38^ ^50^ ^56^ The *organiser* played a key role in setting a professional environment when managing discussions, maintaining confidentiality, and addressing conflicts of interest.^33^ ^55^ Additional responsibilities were setting meeting dates and agendas, and managing time and flow of discussions between committee members.^33^ ^41^ ^43^ ^46^ ^55^ ^56^ ^59^ ^62–64^

### The facilitator

In managing the committee members’ interactions, ^33^ ^34^ ^38^ ^43^ ^46^ ^55^ ^56^ ^59^ the role of *facilitator* was central to preparing consensus decision committee members for their consensus decision committee work. The *facilitator* set ground rules, provided necessary background information to set the context for discussion, and ensured consensus decision committee members had sufficient information to understand the task of the committee and to foster productive discussion.^33^ ^46^ ^48^ To support a balanced discussion, the *facilitator* was responsible for delivering a psychologically safe and inclusive environment, where all committee members could participate and express their views.^34^ ^38^ ^55^ The *facilitator* ensured the consensus meeting adhered to standard procedures, and fostered a collegial atmosphere in the meeting, recognising and addressing challenges that arose from interpersonal conflicts between committee members.^43^ ^48^ ^50^ ^55^ ^56^ ^64^

### The adjudicator

Ensuring balance and fairness in meeting discussions was the goal of the multifaceted adjudicator role.^46^ ^48^ ^52^ ^56^ ^59^ The main focus of the *adjudicator* was on the process of reaching a consensus decision.^40^ ^52^ ^55^ ^56^ The *adjudicator* sometimes guided committee members through regulatory processes or evaluating information, ensuring that the scientific and technical merit of materials presented to the consensus decision committee were discussed fulsomely.^33^ ^34^ ^50^ ^55^ ^56^ The *adjudicator* played a crucial role in consensus decision making—leading and moderating discussions based on objective and justifiable criteria, identifying where committee members agreed and disagreed, bringing confluent points together, recognising and limiting the influence of bias, and directing deliberations towards a decision (i.e., consensus) before bringing discussions among consensus decision committee members to a close, and declaring the consensus-based decision.^33^ ^35^ ^40^ ^44^ ^48^ ^52^ ^55^ ^56^ ^61^ The *adjudicator* fostered an environment for respectful debate by impartially managing conflict, ensuring deliberations stayed on topic, and mediating discrepant opinions.^34^ ^38^ ^55^ ^56^ In grant peer review, the *adjudicator* may also monitor the quality of peer-review and support communication of recommendations and/required feedback.38 51 55 56 63

### The administrator

Governance and record-keeping in the meeting was the domain of the administrator.^55^ ^56^ ^61^ To ensure reliability and accuracy in communication, the *administrator* was accountable for recording detailed meeting notes, summarising discussions, and flagging any potential issues related to ethics, budgets, etc.^55^ ^56^ The administrator also played a support role; they acted as a back-up to the primary consensus decision committee leader if the person was unavailable, and assisted with managerial tasks including documenting and helping to manage conflicts of interests.^55^ ^56^

### Characteristics of consensus decision committee leaders

We extracted 34 statements that reported attributes and characteristics of consensus decision committee leaders. Leaders had relevant experience and knowledge of the field (e.g., in research funding), plus evidence of an active career demonstrated through research or clinical impact.^37^ ^41^ ^49^ ^50^ ^64^ To have success in the facilitator role, leaders needed to prioritise a respectful, inclusive, and collegial approach, and communicate well.^41^ ^42^ ^68^ ^70^ Managerial, organizational, and leadership skills helped leaders facilitate efficient and effective meetings.^33^ ^41^ To support effective decisions, leaders required impartiality and to act decisively.^33^ ^37^ ^41^

We did not synthesise performance indicators reflecting accountability, tenure, time commitment, compensation, and conflict of interest, as the Canadian Institutes of Health Research was the only information source we identified for these elements.

## Discussion

We identified 40 sources reporting the roles and responsibilities of leaders of consensus-based decision meetings. The roles and responsibilities described in our scoping review are a synthesis of research and governance literature from the fields of health sciences, biomedicine, education, psychology, management research, law, ethics, policy and economics. Our review highlights that the field of study of roles and responsibilities of leaders of consensus-based decision meetings is under-developed. Evaluating the process of consensus decision-making has often been limited because of its complexity - influenced by varying factors such as social, organizational, people, or system context ^35^ ^71^. However, given the prominence of consensus committees in peer review (for academic journals and grants) and health guideline development we argue for better defining the roles and responsibilities of committee leaders, train leaders to assume the responsibilities, and evaluating their performance.

Our review uncovered substantial overlap between the roles and responsibilities of the leader (e.g., Chair), and co-leader (e.g., Scientific Officer) of consensus-based decision committees, with the exception of the role of *administrator*, which the co-leader most often assumed. In grant peer-review, as the Chair’s main roles focus on facilitating the peer review process, the Scientific Officer assumed an additional administrative role, capturing key elements of the scientific discussion as a record and to provide a written record of the consensus process and explanation of the consensus decision.

The unique roles we extracted for leaders of consensus-based decision meetings encompassed 3 broad categories: (1) *organizer and/or resource manager*, (2) *facilitator*, and (3) *adjudicator*. An essential leadership quality that we observed across all roles was in facilitating discussion and decisions without dominating the group. The essential role of the leader or chair simultaneously requires balancing (i) dominating the group to counter ‘group think’ or vocal individuals, *and* (ii) relating to each member of the committee as an equal^33^. In this paradox, we argue that exemplary leaders likely leverage their highly-tuned emotional intelligence. Consensus committee leaders can influence meetings in negative and positive ways ^52^. For example, the outcome of the meeting may be associated with the order in which panel members ask questions, a sequence that is set by the committee chair ^34^. Gender differences in the ways that questions are asked may also play a role. ^32^.

There is no standard best practice guide for leading consensus committees. We believe a cogent next move is developing a guideline for executives leading consensus-based meetings. Possible objectives could be to: 1) develop a consensus-based list of roles and responsibilities for leaders, 2) develop educational training to aid development of the necessary skills and 3) create an evaluation framework for leaders’ performance. Committee leaders need advanced skills in facilitating, or at least have access to training to develop those skills ^45^. In 2006, the World Health Organization reviewed the NICE guideline development programme, and proposed recommendations for developing standard training for chairs ^70^. Unfortunately, it is unclear whether or how the recommendations were implemented.

Researchers frequently find themselves serving in roles for which they lack formal training.^6^ One example is journal peer review. Despite consensus among researchers regarding the necessity of training for peer reviewers, very few report receiving any training in peer review ^72^. The leaders of consensus decision processes, including peer review, similarly report an absence of training.

Consequently, there is a need to develop new tailored training and/or supplement existing training to nurture the leadership qualities required for effectively carrying out these roles. We are, sadly, not optimistic of rapid change, given that service as chair is typically voluntary.^58^ Requiring leaders to find additional time for training may simply raise additional barriers. We suggest that organisations who are recruiting consensus committee leaders (and committee members) consider how they will support their people to function effectively in their roles and deliver quality consensus.

### Limitations

We limited our search to works published since 2002 to capture contemporary approaches to the work of consensus committees. For feasibility reasons, we made the pragmatic choice to search databases and web sources for articles and grey literature in English. We included policy, mandates, and guidance documents published in English only, and only searched the websites of only the top 50 international biomedical research funders. Therefore, it is possible that we missed relevant guidance from some international organisations, and our findings likely mirror the cultural contexts of high-income, Western English-speaking countries.

## Conclusion

We described and defined the roles and responsibilities of consensus decision committee leaders. Executives with leadership roles in consensus decision committees, assumed the roles of organiser, facilitator, adjudicator and administrator. Our findings provide a first step in defining how organisations that are responsible for consensus processes might approach training and evaluating the performance of consensus committee leaders.

## Data Availability

The data produced are available online at the Open Science Framework (DOI 10.17605/OSF.IO/K4MC7)

## References

1. Li SA, Yousefi-Nooraie R, Guyatt G, et al. A few panel members dominated guideline development meeting discussions: Social network analysis. J Clin Epidemiol 2022;141:1–10.

2. Fitch K, Bernstein SJ, Aguilar MD, et al. The RAND/UCLA Appropriateness Method User’s Manual: RAND 2001:46.

3. Blazey P, Crossley KM, Ardern CL, et al. It is time for consensus on ‘consensus statements’. Br J Sports Med 2022;56(6):306–07.

4. Blazey P, Scott A, Ardern CL, et al. Consensus methods in patellofemoral pain: how rigorous are they? A scoping review. Br J Sports Med 2024;58(13):733–44.

5. Lasinsky A, Wrightson JG, Khan H, et al. Biomedical research grant resubmission rates, and factors related to success - a scoping review. BMJ Open 2024;14(11):e089927.

6. Sims Gould J, Lasinsky A, Mota A, et al. Threats to grant peer review: A qualitative study. BMJ Open 2025;in press

7. Wrightson JG, Lasinsky A, Snell RR, et al. What factors are important to the success of resubmitted grant applications in health research? A retrospective study of over 20,000 applications to the Canadian Institutes of Health Research. *medRxiv* 2024 doi: 10.1101/2024.05.29.24308137:2024.05.29.24308137.

8. Gallo SA, Carpenter AS, Irwin D, et al. The validation of peer review through research impact measures and the implications for funding strategies. PLoS One 2014;9(9):e106474.

9. Recio-Saucedo A, Crane K, Meadmore K, et al. What works for peer review and decision-making in research funding: a realist synthesis. Res Integr Peer Rev 2022;7(1):2.

10. Gattrell WT, Logullo P, van Zuuren EJ, et al. ACCORD (ACcurate COnsensus Reporting Document): A reporting guideline for consensus methods in biomedicine developed via a modified Delphi. PLoS Med 2024;21(1):e1004326.

11. Guthrie S, Ghiga I, Wooding S. What do we know about grant peer review in the health sciences? An updated review of the literature and six case studies. Santa Monica, CA: RAND Corporation 2018.

12. Witteman HO, Hendricks M, Straus S, et al. Are gender gaps due to evaluations of the applicant or the science? A natural experiment at a national funding agency. The Lancet 2019;393(10171):531–40.

13. van Zuuren EJ, Logullo P, Price A, et al. Existing guidance on reporting of consensus methodology: a systematic review to inform ACCORD guideline development. BMJ Open 2022;12(9):e065154.

14. Gallo SA, Schmaling KB, Thompson LA, et al. Grant reviewer perceptions of the quality, effectiveness, and influence of panel discussion. Res Integr Peer Rev 2020;5:7.

15. European Science Foundation. ESF Survey Analysis Report on Peer Review Practices European Science Foundation2011 [Available from: https://www.esf.org/fileadmin/user_upload/esf/PeerReview-Practices_Survey2011.pdf; accessed September 22 2022.

16. Braun V, Clarke V. One size fits all? What counts as quality practice in (reflexive) thematic analysis. Qualitative Research in Psychology 2021;18(3):328–52.

17. JBI Scoping review network [Available from: https://jbi.global/scoping-review-network/resources.

18. Arksey H, O’Malley L. Scoping studies: towards a methodological framework. Int J Soc Res Methodol 2005;8:19–32.

19. Levac D, Colquhoun H, O’Brien KK. Scoping studies: advancing the methodology. Implement Sci 2010;5:69.

20. Ghannad M, Giustini D, Lasinsky A, et al. Roles and responsibilities when leading consensus meetings OSF 10.17605/OSF.IO/AXMJZ2023 [doi: 10.17605/OSF.IO/AXMJZ.

21. Tricco AC, Lillie E, Zarin W, et al. PRISMA Extension for Scoping Reviews (PRISMA-ScR): Checklist and Explanation. Ann Intern Med 2018;169(7):467–73.

22. Haug C. What Is Consensus and How Is It Achieved in Meetings?: Four Types of Consensus Decision Making. In: Allen JA, Lehmann-Willenbrock N, Rogelberg SG, eds. The Cambridge Handbook of Meeting Science. Cambridge: Cambridge University Press 2015:556–84.

23. Fink A, Kosecoff J, Chassin M, et al. Consensus methods: characteristics and guidelines for use. Am J Public Health 1984;74(9):979–83.

24. Cobo E, Cortes J, Ribera JM, et al. Effect of using reporting guidelines during peer review on quality of final manuscripts submitted to a biomedical journal: masked randomised trial. BMJ 2011;343:d6783.

25. Susskind LE, McKearnen S, Thomas-Lamar J. The consensus building handbook: A comprehensive guide to reaching agreement: Sage publications 1999.

26. Bramer WM, Giustini D, de Jonge GB, et al. De-duplication of database search results for systematic reviews in EndNote. J Med Libr Assoc 2016;104(3):240–3.

27. Braun V, Clarke V. Using thematic analysis in psychology. Qualitative Research in Psychology 2006;3(2):77–101.

28. Braun V, Clarke V, Hayfield N, et al. Answers to frequently asked questions about thematic analysis https://cdn.auckland.ac.nz/assets/psych/about/our-research/documents/Answers%20to%20frequently%20asked%20questions%20about%20thematic%20analysis%20April%202019.pdf2019 [accessed 17 October 2024.

29. Byrne D. A worked example of Braun and Clarke’s approach to reflexive thematic analysis. Quality & Quantity 2022;56:1391–412.

30. Braun V, Clarke V. Successful qualitative research: A practical guide for beginners. London: Sage 2013.

31. Braun V, Clarke V. Reflecting on reflexive thematic analysis. *Qualitative Research in Sport*, Exercise and Health 2019;11(4):589–97.

32. Aritz J, Walker R, Cardon P, et al. Discourse of Leadership: The Power of Questions in Organizational Decision Making. International Journal of Business Communication 2017;54(2):161–81.

33. Bezemer PJ, Nicholson G, Pugliese A. The influence of board chairs on director engagement: A case-based exploration of boardroom decision-making. Corporate Governance: An International Review 2018;26(3):219–34.

34. Broniatowski DA. A method for analysis of expert committee decision-making applied to FDA’s Medical Device Panels. 2011;72:3776–76.

35. Brown P, Hashem F, Calnan M. Trust, regulatory processes and NICE decision-making: Appraising cost-effectiveness models through appraising people and systems. Social Studies of Science 2016;46(1):87–111.

36. Chappell Jr HW, McGregor RR, Vermilyea T. Majority rule, consensus building, and the power of the chairman: Arthur burns and the FOMC. *Journal of Money*, Credit and Banking 2004;36(3 I):407–22.

37. Daykin A, Selman LE, Cramer H, et al. What are the roles and valued attributes of a Trial Steering Committee? Ethnographic study of eight clinical trials facing challenges. Trials 2016;17(1):307.

38. Derrick G, Samuel G. The future of societal impact assessment using peer review: pre-evaluation training, consensus building and inter-reviewer reliability. Palgrave Communications 2017;3(1):17040.

39. Dew K, Stubbe M, Signal L, et al. Cancer care decision making in multidisciplinary meetings. Qualitative Health Research 2015;25(3):397–407.

40. Eklund P, Rusinowska A, De Swart H. Consensus reaching in committees. European Journal of Operational Research 2007;178(1):185–93.

41. Fradgley EA, Booth K, Paul C, et al. Facilitating High Quality Cancer Care: A Qualitative Study of Australian Chairpersons’ Perspectives on Multidisciplinary Team Meetings. J Multidiscip Healthc 2021;14:3429–39.

42. Hakkola L, Dyer SJV. Role conflict: How search committee chairs negotiate faculty status, diversity, and equity in faculty searches. Journal of Diversity in Higher Education 2022;15(5):583–95.

43. Jalil R, Soukup T, Akhter W, et al. Quality of leadership in multidisciplinary cancer tumor boards: development and evaluation of a leadership assessment instrument (ATLAS). World Journal of Urology 2018;36(7):1031–38.

44. Lewandowska K, Smolarska Z. Artistic quality and consensus decision-making: On reviewing panels in the performing arts. International Journal of Media and Cultural Politics 2020;16(2):159–74.

45. Margerum RD. Collaborative Planning:Building Consensus and Building a Distinct Model for Practice. Journal of Planning Education and Research 2002;21(3):237–53.

46. Martin D, Abelson J, Singer P. Participation in health care priority-setting through the eyes of the participants. Journal of Health Services Research and Policy 2002;7(4):222–29.

47. Meyer SR. An analysis of university academic department chairpersons’ resource management decisions. 2017;78

48. Olbrecht M, Bornmann L. Panel peer review of grant applications: what do we know from research in social psychology on judgment and decision-making in groups? Research Evaluation 2010;19(4):293–304.

49. Rittenbach K, Horne CG, O’Riordan T, et al. Engaging people with lived experience in the grant review process. BMC Med Ethics 2019;20(1):95.

50. Selker HP, Buse JB, Califf RM, et al. CTSA Consortium Consensus Scientific Review Committee (SRC) Working Group Report on the SRC Processes. Clinical and Translational Science 2015;8(6):623–31.

51. Taylor C, Atkins L, Richardson A, et al. Measuring the quality of MDT working: an observational approach. BMC Cancer 2012;12(1):202.

52. Wodak R, Kwon W, Clarke I. ’getting people on board’: Discursive leadership for consensus building in team meetings. Discourse and Society 2011;22(5):592–644.

53. Lamprell K, Chittajallu R, Arnolda G, et al. Multidisciplinary team meeting Chairs’ attitudes and perceived facilitators, barriers and ideal improvements to meeting functionality: A qualitative study. Asia Pac J Clin Oncol 2024;20(4):537–45.

54. Roberts V, Carter P, Barnett P, et al. Committee experiences of using formal consensus in healthcare guidelines: a longitudinal qualitative study. BMC Med Inform Decis Mak 2023;23(1):147.

55. Canadian Institutes of Health Research (CIHR). [Available from: https://cihr-irsc.gc.ca/; accessed March 21 2023.

56. National Institutes of Health (NIH). [Available from: https://grants.nih.gov/; accessed March 21 2023.

57. Canadian Cancer Society. Advisory Council on Research (ACOR) 2022 [Available from: https://cancer.ca/en/research/for-researchers/committees/acor; accessed March 21 2023.

58. Congressionally Directed Medical Research Programs. Evaluation of the Congressionally Directed Medical Research Programs Review Process. 2016 [Available from: https://www.ncbi.nlm.nih.gov/books/NBK424516/; accessed March 21 2023.

59. US Food and Drug Administration. FDA Advisory Committees 2020 [Available from: https://www.fda.gov/patients/about-office-patient-affairs/learn-about-fda-advisory-committees#:~:text=The%20role%20of%20the%20chairperson,the%20FDA%20and%20the%20sponsor.; accessed March 21 2023.

60. US Agency for Healthcare Research and Quality. Funding & Grants: Overview – Peer Review Process 2021 [Available from: https://www.ahrq.gov/funding/process/review/peerproc.html; accessed March 21 2023.

61. Engineering and Physical Sciences Research Council. NIHR Academy Selection Committee Chair Role Description 2021 [Available from: https://www.nihr.ac.uk/documents/research-governance-guidelines/12154; accessed March 21 2023.

62. Robert Wood Johnson Foundation. Nominating and Governance Committee Charter 2018 [Available from: https://www.rwjf.org/en/about-rwjf/how-we-work/organizational-policies/nominating-and-governance-committee.html; accessed March 21 2023.

63. Fund for Scientific Research - Flanders. Regulations of the Research Foundation – Flanders governing the internal and external peer review 2018 [Available from: https://www.fwo.be/en/the-fwo/organisation/fwo-expertpanels/regulations-fwo-internal-and-external-peer-review/; accessed March 21 2023.

64. National Institute for Health and Care Research. Role of panel meetings in peer review 2022 [Available from: https://www.ukri.org/councils/epsrc/guidance-for-reviewers/peer-review-panels/role-of-panel-meetings-in-peer-review/; accessed March 21 2023.

65. National Aeronautics and Space Administration. Science Mission Directorate Policy: requirements for research & analysis peer review and selection processes 2022 [Available from: https://science.nasa.gov/science-red/s3fs-public/atoms/files/SPD-22B_peer_review_and_selection_processes101522.pdf/; accessed March 21 2023.

66. US Agency for International Development. USAID Scientific Research Policy 2014; cited 2023 March 21]. Available from: https://pdf.usaid.gov/pdf_docs/PBAAD895.pdf.

67. Western Ontario Health Team. Consensus decision-making process 2020 [Available from: https://lmprimarycare.ca/wp-content/uploads/2020/11/consensus-decision-making-process_oct-15-2020.pdf; accessed March 21 2023.

68. World Health Organization (WHO). Decision-making for guideline development at WHO 2014 [Available from: https://iris.who.int/bitstream/handle/10665/145714/9789241548960_chap16_eng.pdf; accessed March 21 2023.

69. EDUCAUSE. Volunteer Guidelines and Responsibilities [Available from: https://www.educause.edu/about/mission-and-organization/governance-and-leadership/member-committees/volunteer-guidelines-and-responsibilities; accessed March 21 2023.

70. National Institute for Health and Care Excellence (NICE). [Available from: https://www.nice.org.uk/; accessed March 21 2023.

71. Perret C, Powers ST. An investigation of the role of leadership in consensus decision-making. J Theor Biol 2022;543:111094.

72. Willis JV, Ramos J, Cobey KD, et al. Knowledge and motivations of training in peer review: An international cross-sectional survey. PLoS One 2023;18(7):e0287660.

