## Appendix 1 for "What is expected of people who lead meetings where the goal is to reach consensus? A scoping review with implications for improving the quality of health research grant peer review and clinical guideline development"

**Appendix 1. Bibliograhic database search strategies**

**Embase (Ovid) Search**

Search Strategy:

--------------------------------------------------------------------------------

1	consensus/ (93378)

2	consensu*.ti,ab. (277837)

3	exp *decision making/ (92275)

4	"decisionmaking".ti,ab. (4472)

5	"decision making".ti,ab. (240847)

6	Delphi study/ (15402)

7	or/1-6 (578590)

8	chair*.ti. (6004)

9	leadership/ (85130)

10	leader*.ti. (27599)

11	(performance adj5 committee*).ti,ab. (365)

12	(performance adj5 leader*).ti,ab. (1177)

13	(performance adj5 chair*).ti,ab. (659)

14	(role* adj5 committee*).ti,ab. (1280)

15	(role* adj5 leader*).ti,ab. (8307)

16	(role* adj5 chair*).ti,ab. (396)

17	(responsibilit* adj5 committee*).ti,ab. (334)

18	(responsibilit* adj5 leader*).ti,ab. (838)

19	(responsibilit* adj5 chair*).ti,ab. (45)

20	(structure adj5 committee*).ti,ab. (376)

21	(structure adj5 leader*).ti,ab. (908)

22	(structure adj5 chair*).ti,ab. (123)

23	or/8-22 (103629)

24	panel*.mp. (289951)

25	committee*.mp. (187017)

26	meeting*.mp. (233165)

27	grant*.mp. (68034)

28	funding.mp. (139604)

29	"peer review"/ (36058)

30	"peer review".mp. (41494)

31	or/24-30 (891410)

32	7 and 23 and 31 (1562)
