## Appendix 2 for "What is expected of people who lead meetings where the goal is to reach consensus? A scoping review with implications for improving the quality of health research grant peer review and clinical guideline development"

**Appendix 2. Grey literature searches**

| **Grant funding organizations & Foundations** | **Website** | **Country** |
| --- | --- | --- |
| National Research Council Scientific and Technical | https://www.conicet.gov.ar/ | Argentina |
| National Health and Medical Research Council | <https://www.nhmrc.gov.au/> | Australia |
| Fund for Scientific Research – Flanders ^63^ | https://www.fwo.be/en/the-fwo/organisation/fwo-expertpanels/regulations-fwo-internal-and-external-peer-review/ | Belgium |
| Sao Paulo Research Foundation | <https://fapesp.br/en/about> | Brazil |
| Canadian Cancer Society ^57^ | <http://cancer.ca/research/grants%20and%20awards/grants%20to%20individuals.aspx?sc_lang=en> | Canada |
| Canadian Institutes of Health Research ^55^ | <https://cihr-irsc.gc.ca/e/52544.html>  <https://cihr-irsc.gc.ca/e/49564.html>  <https://cihr-irsc.gc.ca/e/49808.html>  Environmental Scan - Potential areas of evaluation for Chairs (Report)  Environmental Scan - Potential areas of evaluation for Scientific Officers (Report) | Canada |
| CHEO Research Institute | <https://www.haloresearch.ca/grants/> | Canada |
| Dairy Farmers of Canada | [https://www.dairynutrition.ca](https://www.dairynutrition.ca/) | Canada |
| Health Research BC | <https://www.msfhr.org/> | Canada |
| Heart and Stroke Foundation of Canada | <https://www.heartandstroke.ca/search-results-page?q=chair+roles> | Canada |
| Multiple Sclerosis Society of Canada | <http://www.mssociety.ca/en/research/researchfunding.htm> | Canada |
| Sick Kids Foundation | <http://www.sickkidsfoundation.com/grants/knowledge.asp> | Canada |
| The Lawson Foundation | <http://www.lawson.ca/index.php?option=com_content&task=view&id=12&Itemid=26> | Canada |
| Chinese Academy of Sciences | <https://english.cas.cn/> | China |
| Chinese Center for Disease Control and Prevention | <https://www.chinacdc.cn/en/> | China |
| Chinese Government Ministry of Health | <http://en.nhc.gov.cn/> | China |
| The National Administration of Traditional Chinese Medicine | <http://satcm.gov.cn/> | China |
| The National Natural Science Foundation of China | <https://www.nsfc.gov.cn/english/site_1/index.html> | China |
| Academy of Finland | <https://www.aka.fi/en/> | Finland |
| French National Cancer Institute | <https://en.e-cancer.fr/> | France |
| French National Research Agency | <https://anr.fr/en/> | France |
| Institut Pasteur | <https://research.pasteur.fr/en/#_ga=2.242556737.251870569.1660168508-1451756891.1660168508> | France |
| National Agency for AIDS Research | <https://www.anrs.fr/en> | France |
| National Agency for the Evaluation of Research and Higher Education | [https://www.aeres-evaluation.fr](https://www.aeres-evaluation.fr/) | France |
| National Centre for Scientific Research | <https://www.cnrs.fr/en/cnrs> | France |
| National Institute of Health and Medical Research | <https://www.inserm.fr/en/home/> | France |
| Federal Ministry of Education and Research | <https://www.bmbf.de/bmbf/en/home/home_node.html> | Germany |
| Federal Ministry of Health | <https://www.bundesgesundheitsministerium.de/en/> | Germany |
| German Cancer Consortium | <https://dktk.dkfz.de/en> | Germany |
| German Center for Cardiovascular Research | <https://dzhk.de/en/> | Germany |
| German Center for Diabetes Research | <https://www.dzd-ev.de/en/the-dzd/index.html> | Germany |
| German Center for Infection Research | <https://www.dzif.de/en> | Germany |
| German Centre for Lung Research | <https://dzl.de/en/> | Germany |
| German Centre for Neurodegenerative Diseases | <https://www.dzne.de/en/> | Germany |
| German Research Foundation | <https://www.dfg.de/en/research_funding/index.html> | Germany |
| Indian Council of Medical Research | <https://www.icmr.gov.in/> | India |
| Italian National Institute of Health | <https://www.iss.it/web/iss-en> | Italy |
| Ministry of Health Italy | <https://moh-it.pure.elsevier.com/> | Italy |
| Japan Science and Technology Agency | <https://www.jst.go.jp/EN/programs/funding.html> | Japan |
| Japan Society for the Promotion of Science | <https://www.jsps.go.jp/english/> | Japan |
| The Netherlands Organisation for Health Research and Development | <https://www.zonmw.nl/en/> | Netherlands |
| Health Research Council of New Zealand | <https://www.hrc.govt.nz/grants-funding/funding-opportunities> | New Zealand |
| Research Council of Norway | <https://www.forskningsradet.no/en/> | Norway |
| Foundation Dam | <https://dam.no/> | Norway |
| Singapore National Medical Research Council | <https://www.nmrc.gov.sg/> | Singapore |
| Korean National Research Foundation | <https://www.nrf.re.kr/eng/main/> | South Korea |
| National Institute of Health | <https://nih.go.kr/eng/> | South Korea |
| Spanish National Cancer Centre | <https://www.cnio.es/en/> | Spain |
| Spanish National Research Council | <https://www.csic.es/en/csic> | Spain |
| Health Research Fund | No homepage found | Spain |
| Canary Islands Foundation for Research and Health | <https://fciisc.org/> | Spain |
| Institute of Biomedicine of Valencia | <https://www.csic.es/en/investigation/institutes-centres-units/institute-biomedicine-valencia> | Spain |
| Biomedicine Institute of Seville | <https://www.ibis-sevilla.es/> | Spain |
| Carlos III Health Institute | <https://portalfis.isciii.es/es/Paginas/inicio.aspx> | Spain |
| Spanish National Health System | <https://www.sanidad.gob.es/en/organizacion/sns/libroSNS.htm> | Spain |
| Swedish Research Council | <https://www.vr.se/english.html> | Sweden |
| Swiss National Science Foundation | <https://www.snf.ch/en> | Switzerland |
| British Heart Foundation | <https://www.bhf.org.uk/for-professionals/information-for-researchers/how-to-apply> | United Kingdom |
| Cancer Research UK | <https://www.cancerresearchuk.org/funding-for-researchers/our-funding-schemes> | United Kingdom |
| Engineering and Physical Sciences Research Council ^61^ | <https://www.nihr.ac.uk/documents/research-governance-guidelines/12154> | United Kingdom |
| National Institute for Health and Care Research ^64^ | <https://www.ukri.org/councils/epsrc/guidance-for-reviewers/peer-review-panels/role-of-panel-meetings-in-peer-review/> | United Kingdom |
| UK Medical Research Council | <https://www.ukri.org/councils/mrc/> | United Kingdom |
| Wellcome Trust | <https://wellcome.org/grant-funding> | United Kingdom |
| Alfred P. Sloan Foundation | <https://sloan.org/> | United States |
| American Cancer Society | <https://www.cancer.org/research/we-fund-cancer-research/apply-research-grant.html> | United States |
| American Heart Association | <https://professional.heart.org/en/research-programs/application-information#resources> | United States |
| Bill and Melinda Gates Foundation | <https://www.gatesfoundation.org/about/how-we-work/grant-opportunities> | United States |
| Breast Cancer Research Foundation | <https://www.bcrf.org/> | United States |
| Burroughs Wellcome Fund | [https://www.bwfund.org](https://www.bwfund.org/) | United States |
| Centers for Disease Control and Prevention | <https://www.cdc.gov/grants/index.html> | United States |
| Congressionally Directed Medical Research Programs ^58^ | <https://www.ncbi.nlm.nih.gov/books/NBK424516/> | United States |
| Crohn’s & Colitis Foundation | [https://www.crohnscolitisfoundation.org](https://www.crohnscolitisfoundation.org/) | United States |
| Department of Defense | <https://basicresearch.defense.gov/> | United States |
| Department of Veterans Affairs | <https://www.research.va.gov/> | United States |
| Flinn Foundation | <https://flinn.org/> | United States |
| Foundation for Physical Therapy Research | [https://foundation4pt.org](https://foundation4pt.org/) | United States |
| Gordon & Betty Moore Foundation | <https://www.moore.org/home> | United States |
| Hewlett Foundation | [https://hewlett.org](https://hewlett.org/) | United States |
| Howard Hughes Medical Institute | https://www.hhmi.org/programs/open-competitions | United States |
| Kenneth Rainin Foundation | <https://krfoundation.org/> | United States |
| National Aeronautics and Space Administration ^65^ | [https://science.nasa.gov/science-red/s3fs-public/atoms/files/SPD-22B_peer_review_and_selection_processes101522.pdf /](https://science.nasa.gov/science-red/s3fs-public/atoms/files/SPD-22B_peer_review_and_selection_processes101522.pdf%20/) | United States |
| National Institutes of Health (NIH) ^56^ | <https://grants.nih.gov/grants/peer/guidelines_general/Chair_orientation.pdf>  <https://grants.nih.gov/grants/peer-review.htm> | United States |
| Patient Centered Outcomes Research Institute | <https://www.pcori.org/funding-opportunities> | United States |
| Robert Wood Johnson Foundation ^62^ | <https://www.rwjf.org/en/about-rwjf/how-we-work/organizational-policies/nominating-and-governance-committee.html> | United States |
| Templeton World | [https://www.templetonworldcharity.org](https://www.templetonworldcharity.org/) | United States |
| US Agency for Healthcare Research and Quality ^60^ | <https://www.ahrq.gov/funding/process/review/peerproc.html> | United States |
| US Agency for International Development ^66^ | <https://pdf.usaid.gov/pdf_docs/PBAAD895.pdf> | United States |
| US Department of Agriculture | <https://www.nifa.usda.gov/grants/funding-opportunities> | United States |
| US Department of Defense | <https://www.grants.gov/learn-grants/grant-making-agencies/department-of-defense.html> | United States |
| US Department of Health and Human Services | <https://www.grants.gov/web/grants/learn-grants/grant-making-agencies/department-of-health-and-human-services.html> | United States |
| US Environmental Protection Agency | <https://www.epa.gov/grants> | United States |
| US Food and Drug Administration ^59^ | <https://www.fda.gov/patients/about-office-patient-affairs/learn-about-fda-advisory-committees#:~:text=The%20role%20of%20the%20chairperson,the%20FDA%20and%20the%20sponsor>. | United States |
| William T. Grant Foundation | <https://wtgrantfoundation.org/grants/research-grants-improving-use-research-evidence> | United States |
| European Commission | <https://ec.europa.eu/info/research-and-innovation/funding/funding-opportunities/funding-programmes-and-open-calls_en> | European Union |
| European Research Council | <https://erc.europa.eu/> | European Union |
| Innovative Medicines Initiative | <https://www.imi.europa.eu/> | European Union |
| **Other Organizations**  **(Health, Government and Business)** | **Website** | **Country** |
| Government of Canada | https://www.canada.ca/en/treasury-board-secretariat/services/professional-development/key-leadership-competency-profile/examples-effective-ineffective-behaviours.html | Canada |
| House of Commons | <https://www.ourcommons.ca/procedure/procedure-and-practice-3/ch_20_6-e.html> | Canada |
| Western Ontario Health Team ^67^ | https://lmprimarycare.ca/wp-content/uploads/2020/11/consensus-decision-making-process_oct-15-2020.pdf | Canada |
| Ministry of Agriculture, Food and Rural Affairs | <http://www.omafra.gov.on.ca/english/nfporgs/16-005.pdf> | Canada |
| National Institute for Health and Care Excellence (NICE) ^70^ | <https://www.nice.org.uk/process/pmg20/chapter/decision-making-committees>  <https://g-i-n.net/wp-content/uploads/2021/08/Role-of-chairs-in-supporting-PPI.pdf> | United Kingdom |
| EDUCAUSE ^69^ | <https://www.educause.edu/about/mission-and-organization/governance-and-leadership/member-committees/committee-guidelines-and-responsibilities> | United States |
| Institute for Local Government | http://www.ca-ilg.org/sites/main/files/file-attachments/understanding_the_role_of_chair_nov_2012_1.pdf | United States |
| World Health Organisation ^68^ | <https://apps.who.int/iris/bitstream/handle/10665/145714/9789241548960_chap16_eng.pdf> | International |
| Diligent | https://www.diligent.com/insights/roles-responsibilities/what-is-the-role-of-a-committee-chair/ | International |
