## Appendix 3 for "What is expected of people who lead meetings where the goal is to reach consensus? A scoping review with implications for improving the quality of health research grant peer review and clinical guideline development"

**Appendix 3. Articles excluded at full text screening stage, with reasons**

Not about Chair roles/responsibilities

1. INTERNATIONAL PEER REVIEW EXPERT PANEL REPORT. 2017

2. Albert M, Laberge S. Confined to a tokenistic status: Social scientists in leadership roles in a national health research funding agency. *Social Science & Medicine* 2017;185:137-46.

3. Alfandari R. Legal Advocacy for Parents in Child Protection: Not a Question of If, but a Question of How. *British Journal of Social Work* 2019;49(6):1601-18.

4. Allen P, Parks RG, Kang SJ, Dekker D, Jacob RR, Mazzucca-Ragan S, Brownson RC. Practices Among Local Public Health Agencies to Support Evidence-Based Decision Making: A Qualitative Study. *Journal of public health management and practice : JPHMP* 2022 doi: https://dx.doi.org/10.1097/PHH.0000000000001653

5. Almost J, Wolff AC, Stewart‐Pyne A, McCormick LG, Strachan D, D'Souza C. Managing and mitigating conflict in healthcare teams: an integrative review. *Journal of Advanced Nursing (John Wiley & Sons, Inc)* 2016;72(7):1490-505.

6. Aloo PM. Intra-district resource allocation and criteria used for student based funding in Urban school districts. 2014;74

7. Andersen IE, Jaeger B. Scenario workshops and consensus conferences: Towards more democratic decision-making. *Science and Public Policy* 1999;26(5):331-40.

8. Ashkar CE, Nakkash R, Matar A, Makhoul J. Behind the scenes of research ethics committee oversight: a qualitative research study with committee chairs in the Middle East and North Africa region. *BMC medical ethics* 2024;25(1):86.

9. Aulisio MP, Arnold RM. Role of the ethics committee: Helping to address value conflicts or uncertainties. *Chest* 2008;134(2):417-24.

10. Austen-Smith D, Feddersen TJ. Information aggregation and communication in committees. *Philosophical Transactions of the Royal Society B: Biological Sciences* 2009;364(1518):763-69.

11. Avdagovska M, Stafinski T, Ballermann M, Menon D, Olson K, Paul P. Tracing the decisions that shaped the development of MyChart, an electronic patient portal in Alberta, Canada: Historical research study. *Journal of Medical Internet Research* 2020;22(5):e17505.

12. Bakerjian D, Beverly C, Burger SG, Carter D, Dornberger S, Eliopoulos C, Remsburg R. Gerontological nursing leadership in the Advancing Excellence Campaign: moving interdisciplinary collaboration forward. *Geriatric nursing (New York, NY)* 2014;35(6):417-22.

13. Beca JP, Guerrero JL. Do not resuscitate orders for pediatric patients: The role of a clinical ethics committee in a developing country. *Revista Panamericana de Salud Publica/Pan American Journal of Public Health* 1997;1(2):138-43.

14. Ben Charif A, Croteau J, Adekpedjou R, Zomahoun HTV, Adisso EL, Légaré F. Implementation Research on Shared Decision Making in Primary Care: Inventory of Intracluster Correlation Coefficients. *Medical Decision Making* 2019;39(6):661-72.

15. Ben-Yashar R, Koh WTH, Nitzan S. Is specialization desirable in committee decision making? *Theory and Decision* 2012;72(3):341-57.

16. Bregar A. Application of a hybrid Delphi and aggregation–disaggregation procedure for group decision-making. *EURO Journal on Decision Processes* 2019 doi: https://doi.org/10.1007/s40070-018-0094-3:1.

17. Caillaud B, Tirole J. Consensus Building: How to Persuade a Group. *The American Economic Review* 2007;97(5):1877-900.

18. Campbell G, Sprague KL. The state of drug decision-making: Report on a survey of P & T Committee structure and practices. *Formulary* 2001;36(9):644-55.

19. Chahine S, Cristancho S, Padgett J, Lingard L. How do small groups make decisions? : A theoretical framework to inform the implementation and study of clinical competency committees. *Perspectives on medical education* 2017;6(3):192-98.

20. Chambers FG. Effective Decision-Making and Conflict Resolution for Church Leadership Teams and Governing Boards. Liberty University, 2023.

21. Chatterjee A. Decision-making among philanthropic foundations in the US: Factors that influence international giving. 2018;79

22. Cho HY. An overview of the national immunization policy making process: The role of the Korea expert committee on immunization practices. *Korean Journal of Pediatrics* 2012;55(1):1-5.

23. Choi S, Schnurr S. Exploring distributed leadership: Solving disagreements and negotiating consensus in a 'leaderless' team. *Discourse Studies* 2014;16(1):3-24.

24. Chong PS, Benli ÖS. Consensus in team decision making involving resource allocation. *Management Decision* 2005;43(9):1147-60.

25. D'Aunno T, Alexander JA, Lan J. Creating value for participants in multistakeholder alliances: The shifting importance of leadership and collaborative decision-making over time. *Health Care Management Review* 2017;42(2):100-11.

26. Day DV, Sessa VI. Accounting for choice: How committees justify executive selection decisions. *The Psychologist-Manager Journal* 2003;6(2):79-95.

27. Eisenberg T, Fisher T, Rosen-Zvi I. Group decision making on appellate panels: Presiding justice and opinion justice influence in the Israel Supreme Court. *Psychology, Public Policy, and Law* 2013;19(3):282-96.

28. Fleming AS. Group decision-making and leadership: An experimental examination in an executive compensation scenario. *Advances in Management Accounting* 2008;17:113-49.

29. Floress K, Prokopy LS, Ayres J. Who's in charge: Role clarity in a Midwestern watershed group. *Environmental Management* 2011;48(4):825-34.

30. Foglia MB, Pearlman RA, Bottrell M, Altemose JK, Fox E. Ethical challenges within Veterans Administration healthcare facilities: perspectives of managers, clinicians, patients, and ethics committee chairpersons. *American Journal of Bioethics* 2009;9(4):28-36.

31. Frank AK, O’Sullivan P, Mills LM, Muller-Juge V, Hauer KE. Clerkship Grading Committees: the Impact of Group Decision-Making for Clerkship Grading. *Journal of General Internal Medicine* 2019;34(5):669-76.

32. Freely P. Perceptions of the Grant Decision-Making Process: a Study of Foundation Grantmakers and Grant Seekers Who Focus on Youth Violence in Chicago, Illinois. Loyola University Chicago, 2023.

33. Gerling K, Grüner HP, Kiel A, Schulte E. Information acquisition and decision making in committees: A survey. *European Journal of Political Economy* 2005;21(3):563-97.

34. Gessner BD, Duclos P, DeRoeck D, Nelson EAS. Informing decision makers: Experience and process of 15 National Immunization Technical Advisory Groups. *Vaccine* 2010;28(SUPPL. 1):A1-A5.

35. Godos-Díez JL, Cabeza-García L, Alonso-Martínez D, Fernández-Gago R. Factors influencing board of directors’ decision-making process as determinants of CSR engagement. *Review of Managerial Science* 2018;12(1):229-53.

36. Grubb B. Physician Associate (PA) Leaders in Executive, Academic, and Clinical Roles: Consensus Building for a Novel Leadership Framework. Pepperdine University, 2024.

37. Guillemin M, Gillam L, Rosenthal D, Bolitho A. Human research ethics committees: examining their roles and practices. *J Empir Res Hum Res Ethics* 2012;7(3):38-49.

38. Guldbrandsson K, Stenström N, Winzer R. The DECIDE evidence to recommendation framework adapted to the public health field in Sweden. *Health Promotion International* 2016;31(4):749-54.

39. Häge FM. Bureaucrats as law-makers: Committee decision-making in the EU council of ministers. *Bureaucrats as Law-makers: Committee Decision-Making in the EU Council of Ministers* 2012 doi: 10.4324/9780203104507:1-224.

40. Hales B. The process by which three elementary principals implement response to intervention with English learners. 2018;79

41. Halvorsen K, Sarangi S. Team decision-making in workplace meetings: The interplay of activity roles and discourse roles. *Journal of Pragmatics* 2015;76:1-14.

42. Karuga R, Khan S, Kok M, Moraa M, Mbindyo P, Broerse J, Dieleman M. Teamwork in community health committees: a case study in two urban informal settlements. *BMC health services research* 2023;23(1):1373.

43. Kirman CR, Simon TW, Hays SM. Science peer review for the 21st century: Assessing scientific consensus for decision-making while managing conflict of interests, reviewer and process bias. *Regulatory Toxicology and Pharmacology* 2019;103:73-85.

44. Lafrance GA. Stakeholders' views of an ideal Ontario college boards' decision-making roles. 2010;70:3739-39.

45. Lamb B, Payne H, Vincent C, Sevdalis N, Green JSA. The role of oncologists in multidisciplinary cancer teams in the UK: an untapped resource for team leadership? *Journal of Evaluation in Clinical Practice* 2011;17(6):1200-06.

46. Lamb BW, Wong HWL, Vincent C, Green JSA, Sevdalis N. Teamwork and team performance in multidisciplinary cancer teams: development and evaluation of an observational assessment tool. *BMJ Quality & Safety* 2011;20(10):849-56.

47. Lupi A, Suchá D, Cundari G, Fink N, Alkadhi H, Budde RPJ, Caobelli F, De Cecco CN, Galea N, Hrabak-Paar M, Loewe C, Luetkens JA, Muscogiuri G, Natale L, Nikolaou K, Pirnat M, Saba L, Salgado R, Williams MC, Wintersperger BJ, Vliegenthart R, Francone M, Pepe A. Standards for conducting and reporting consensus and recommendation documents: European Society of Cardiovascular Radiology policy from the Guidelines Committee. *Insights into Imaging* 2024;15(1)

48. Maturo A, Ventre AGS. Reaching consensus in multiagent decision making. *International Journal of Intelligent Systems* 2010;25(3):266-73.

49. Nelson S, Abimbola S, Mangubhai S, Jenkins A, Jupiter S, Naivalu K, Naivalulevu V, Negin J. Understanding the decision-making structures, roles and actions of village-level water committees in Fiji. *International Journal of Water Resources Development* 2022;38(3):518-35.

50. Niu L, Zhao R, Wei Y. How does differential leadership affect team decision-making effectiveness? The role of thriving at work and cooperative goal perception. *Chinese Management Studies* 2024;18(1):91-106.

51. Nor NFM, Aziz J. Discourse analysis of decision making episodes in meetings: Politeness theory and critical discourse analysis. *3L: Language, Linguistics, Literature* 2010;16(2):66-92.

52. Page SA, Nyeboer J. Improving the process of research ethics review. *Res Integr Peer Rev* 2017;2:14.

53. Parreiras RO, Ekel PY, Morais DC. Fuzzy Set Based Consensus Schemes for Multicriteria Group Decision making Applied to Strategic Planning. *Group Decision and Negotiation* 2012;21(2):153-83.

54. Patton DE, Olin SS. Scientific Peer Review to Inform Regulatory Decision Making: Leadership Responsibilities and Cautions. *Risk Analysis* 2006;26(1):5-16.

55. Poitras J, Bowen RE. A Framework for Understanding Consensus-Building Initiation. *Negotiation Journal* 2002;18(3):211.

56. Renz MA. Paving consensus: Enacting, challenging, and revising the consensus process in a cohousing community. *Journal of Applied Communication Research* 2006;34(2):163-90.

57. Roman F, Verma H, Jermann P, Dillenbourg P. Group dynamics findings from coordination in problem solving and decision making meetings. *GROUP'12 - Proceedings of the ACM 2012 International Conference on Support Group Work* 2012 doi: 10.1145/2389176.2389232:305-06.

58. Silverman J, Lidz CW, Clayfield J, Murray A, Simon LJ, Maranda L. Factors Influencing IACUC Decision Making: Who Leads the Discussions? *Journal of Empirical Research on Human Research Ethics* 2017;12(4):209-16.

59. Skorupinski B, Baranzke H, Hans Werner I, Meinhardt M. Consensus Conferences - A Case Study: Publiforum in Switzerland with Special Respect to the Role of Lay Persons and Ethics. *Journal of Agricultural and Environmental Ethics* 2007;20(1):37-52.

60. Steiner Davis MLE, Conner TR, Miller-Bains K, Shapard L. What makes an effective grants peer reviewer? An exploratory study of the necessary skills. *PLoS One* 2020;15(5):e0232327.

61. Tamagna VM. How State and National Political Leaders Employ Values-Based Leadership in Service of Passing Policy Initiatives, Building Consensus, and Building Followings. Concordia University Chicago, 2023.

62. van Baalen S, Carusi A, Sabroe I, Kiely DG. A social-technological epistemology of clinical decision-making as mediated by imaging. *Journal of Evaluation in Clinical Practice* 2017;23(5):949-58.

63. Vojtecky MA. Status and control in voluntary community health planning groups. *Medical Care* 1982;20(12):1168-77.

64. Weill D, Benden C, Corris PA, Dark JH, Davis RD, Keshavjee S, Lederer DJ, Mulligan MJ, Patterson GA, Singer LG, Snell GI, Verleden GM, Zamora MR, Glanville AR. A consensus document for the selection of lung transplant candidates: 2014 - An update from the Pulmonary Transplantation Council of the International Society for Heart and Lung Transplantation. *Journal of Heart and Lung Transplantation* 2015;34(1):1-15.

65. Zápal J. Crafting consensus. *Public Choice* 2017;173(1-2):169-200.

66. Zug RLL. Reflections on Leadership and Decision-Making Challenges During the COVID-19 Pandemic: Narratives of Upper Division Heads in Quaker Schools. University of Pennsylvania, 2024.

67. Trent MS. Participatory Leadership: A Qualitative Narrative Study of a Public South-Central Virginia School Division and Their Perceptions of Teachers Participating in the Instructional Decision-Making Process. Marymount University, 2024.

No consensus decisions

68. Al Flaiti SA. Influencers affecting funding decision-making in universal health care organizations in Oman. 2017;78

69. Albarado AR. Two-year college presidents' perceptions of leader attributes that contribute to successfully securing alternative revenue. 2018;78

70. Arms D, Stalter AM. Serving on Organizational Boards: What Nurses Need to Know. *Online Journal of Issues in Nursing* 2016;21(2):1-1.

71. Armstrong G. Leadership in times of change: An examination of a merger experience. 2012;73:2006-06.

72. Bak J, Oh A. Conversational decision-making model for predicting the king's decision in the annals of the joseon dynasty. *Proceedings of the 2018 Conference on Empirical Methods in Natural Language Processing, EMNLP 2018* 2018:956-61.

73. Bauer A, Evans-Lacko S, Knapp M. Valuing recovery-oriented practice at the interface between mental health services and communities: The role of organisational characteristics and environments. *International Journal of Social Psychiatry* 2019;65(2):136-43.

74. Bekemeier B, Chen ALT, Kawakyu N, Yang Y. Local public health resource allocation: Limited choices and strategic decisions. *American Journal of Preventive Medicine* 2013;45(6):769-75.

75. Best S, Williams SJ. What Have We Learnt About the Sourcing of Personal Protective Equipment During Pandemics? Leadership and Management in Healthcare Supply Chain Management: A Scoping Review. *Frontiers in Public Health* 2021;9:765501.

76. Bolster CJ, Quirk B. How CFOs can help compensation committees make better decisions. *Healthcare financial management : journal of the Healthcare Financial Management Association* 2010;64(9):128-32.

77. Bonazza J, Farrell PM, Albanese M, Kindig D. Collaboration and peer review in medical schools' strategic planning. *Academic Medicine* 2000;75(5):409-18.

78. Boyes M, Potter T, Andkjaer S, Lindner M. The role of planning in outdoor adventure decision-making. *Journal of Adventure Education and Outdoor Learning* 2019;19(4):343-57.

79. Braddom-Ritzler C. Moving up: successful negotiation for the position of academic chair. *American Journal of Physical Medicine & Rehabilitation* 2005;84(9):712-18.

80. Charochak SM. Central office leaders' role in supporting principal autonomy and accountability in a turnaround district. 2018;79

81. Dona LH. Academic Competencies for Medical Faculty.

82. Dougherty KL, Kisaalita A, McKissick J, Katz E. Stopping rules for majority voting: A public choice experiment. *Journal of Economic Behavior & Organization* 2020;175:353-64.

83. Edwards DJ. Attributes and Competencies for Hybrid Team Leaders Post-COVID-19: A Delphi Study. Dallas Baptist University, 2023.

84. Edwards-Barrios CL. Executive Leader Enactment of Empathy in Organizational Decision-Making a Multiple-Method Exploratory Sequential Case Study. University of Charleston - Beckley, 2023.

85. Glenn P. Developing a Decision-Making Framework for Leadership Teams. Royal Roads University (Canada), 2023.

86. Gordon SFA. Teachers’ Experiences in the School’s Leadership Decision-Making Processes Within School Districts. Grand Canyon University, 2023.

87. Grossman MP. An evaluation of leadership competencies and professional development recommendations for the leaders of Delaware adult education. 2010;71:422-22.

88. Hancock TM. The business of universities and the role of department chair. *International Journal of Educational Management* 2007;21(4):306-14.

89. Hebert J, Robitaille H, Turcotte S, Legare F. Online Dissemination Strategies of a Canada Research Chair: Overview and Lessons Learned. *JMIR research protocols* 2017;6(2):e27.

90. Hobgood CD, Draucker C. Barriers, Challenges, and Solutions: What Can We Learn About Leadership in Academic Medicine From a Qualitative Study of Emergency Medicine Women Chairs? *Acad Med* 2022;97(11):1656-64.

91. Hoven M, Segers M, Gevers J, Van den Bossche P. Leader airtime management and team effectiveness in emergency management command and control (EMCC) teams. *Ergonomics* 2023;66(10):1565-81.

92. Huizenga S, van de Bovenkamp H, Oldenhof L, Bal R. The clocks run at slightly different speeds. Clashing timeframes in COVID-19 health risk governance. *Health, Risk and Society* 2023;25(7-8):366-86.

93. Jestine P, Gilli K, Knappstein M. Identifying key leadership competencies for digital transformation: evidence from a cross-sectoral Delphi study of global managers. *Leadership & Organization Development Journal* 2023;44(3):392-406.

94. Kanninen TH, Haggman-Laitila A, Tervo-Heikkinen T, Kvist T. Nursing shared governance at hospitals - it's Finnish future? *Leadership in health services (Bradford, England)* 2019;32(4):558-68.

95. Kelley K. Ethical Decision-Making: The Impact of Gender, Personality and Managerial Experience on Leadership. Capella University, 2023.

96. Lawson BP. The influence of individual audit committee chairs, ceos, and cfos on corporate reporting and operating decisions. 2013;74

97. Lindenauer PK, Benjamin EM, Naglieri-Prescod D, Fitzgerald J, Pekow P. The role of the institutional review board in quality improvement: a survey of quality officers, institutional review board chairs, and journal editors. *The American Journal of Medicine* 2002;113(7):575-79.

98. McBride A, Li ST, Turner TL, Vinci RJ. Roles, Responsibilities, and Value of a Vice Chair of Education in Academic Pediatric Departments. *J Pediatr* 2020;217:4-6 e1.

99. McDougal JA, Brooks CM, Albanese M. Achieving consensus on leadership competencies and outcome measures: The Pediatric Pulmonary Centers' experience. *Evaluation & the health professions* 2005;28(4):428-46.

100. Megheirkouni M. Leadership and decision-making styles in large-scale sporting events. *Event Management* 2018;22(5):785-801.

101. Reeder M. Exploring Information Technology Leaders' Decision-making Process While Leading Projects Using a Hybrid Approach: A Qualitative Case Study. American College of Education, 2024.

102. Rennie SM, Prieur L, Platt M. Communication style drives emergent leadership attribution in virtual teams. *Frontiers in psychology* 2023;14(101550902):1095131.

103. Scott R, Hawarden A, Russell B, Edmondson RJ. Decision-Making in Gynaecological Oncology Multidisciplinary Team Meetings: A Cross-Sectional, Observational Study of Ovarian Cancer Cases. *Oncology Research and Treatment* 2020;43(3):70-76.

104. Slowka S. An end to the "muffin meeting": Conceptualizing power and navigating tokenism in patient engagement for health leaders. *Healthcare management forum* 2024;37(4):296-300.

105. Sry R, Tialonawarmi F, Syahmardi Y. The impact of leader power on organizational development: a strategic approach to decision-making. *Verslas : Teorija ir Praktika* 2023;24(2):557-70.

106. Wihl J, Rosell L, Bendahl PO, De Mattos CBR, Kinhult S, Lindell G, von Steyern FV, Nilbert M. Leadership perspectives in multidisciplinary team meetings; observational assessment based on the ATLAS instrument in cancer care. *Cancer Treatment and Research Communications* 2020;25:100231.

Published before 2002

107. Anonymous. Institutional ethics committee's roles, responsibilities, and benefits for physicians. *Minnesota medicine* 1985;68(8):605-12.

108. Bachrach DJ. Developing physician leaders in academic medical centers: Part 1: their changing role. *Medical Group Management Journal* 1996;43(6):35.

109. Bachrach DJ. Developing physician leaders in academic medical centers. *Medical Group Management Journal* 1997;44(1):34-43.

110. Balls HR. Decision‐making: the role of the deputy minister. *Canadian Public Administration* 1976;19(3):417-31.

111. Beil M, Litscher JE. Consensus bargaining in Wisconsin state government: A new approach to labor negotiation. *Public Personnel Management* 1998;27(1):39-48.

112. Bonkovsky FO. Contending medical decision models. *Theoretical medicine and bioethics* 2001;22(3):193-210.

113. Booth DB. The Department Chair: Professional Development and Role Conflict. 1982

114. Carey RG. Selecting leaders through shared decision making. *Hospital Progress* 1979;60(11):53-55.

115. Christine N. The "Legislative Backbone" keeping the Institution upright? The Role of European Parliament Committees in the EU Policy-Making Process. 2001

116. Csikai EL, Sales E. The emerging social work role on hospital ethics committees: a comparison of social worker and chair perspectives. *Social Work* 1998;43(3):233-42.

117. Damusis VB. Small Group Discussion Characteristics Influencing Perception of Group Consensus and Leader Behavior. 1972 (7228418):226.

118. Elmi FN. Factors Affecting Faculty Role Consensus in a Public Two-Year College. 1975 (7601733):213.

119. Emerick RE. The Concept of Consensus: A Study of the Sources of Conceptual Diversity within the Theoretical Consensus Literature. 1971 (7130792):272.

120. English CB. The art of leading meetings. *The American journal of occupational therapy : official publication of the American Occupational Therapy Association* 1987;41(5):321-26.

121. Fagan LA, Walter SM. Building an interdisciplinary team. Strategies for leadership, consensus building, meetings, and performance reviews. *Rehab management* 1998;11(1):24-28.

122. Franklin IM. A consensus statement on unrelated donor bone marrow transplantation from the consensus panel chaired by EC Gordon-Smith. *Bone Marrow Transplantation* 1997;19(10):959-62.

123. Geonetta SC. An Experimental Study of the Relationship of Orientation and Consensus, Cohesiveness, Satisfaction, and Credibility in Leaderless Groups and Groups with Appointed Leaders. 1974 (7501700):133.

124. Godwin WF. Subgroup Pressures in Small-Group Consensus Processes. 1970 (7106855):148.

125. Hafertepe EC. Systemwide board assessment. *Health progress (Saint Louis, Mo)* 1987;68(1):82-86.

126. Hall RL. Participation and Purpose in Committee Decision Making. *American Political Science Review* 1987;81(1):105-27.

127. Heher JR. Chairman's outlook: Monitoring of the procedure whereby applications for medical staff membership and clinical privileges are reviewed. *Trustee : the journal for hospital governing boards* 1975;28(2):10.

128. Ho T, Antunes P. Developing a tool to assist electronic facilitation of decision-making groups. *String Processing and Information Retrieval Symposium and International Workshop on Groupware, SPIRE 1999 and CRIWG 1999* 1999 doi: 10.1109/spire.1999.796601:243-52.

129. Macklin R. Disagreement, consensus, and moral integrity. *Kennedy Institute of Ethics Journal* 1996;6(3):289-311.

130. Martin PA. Bioethics and the whole: pluralism, consensus, and the transmutation of bioethical methods into gold. *The Journal of law, medicine & ethics : a journal of the American Society of Law, Medicine & Ethics* 1999;27(4):316-294.

131. McGregor L. Improving the quality and speed of decision making. *Journal of Change Management* 2001;2(4):344-56.

132. McKerrow W. The roles and responsibilities of the medical staff organization. *Health law in Canada* 1991;11(4):91-93.

133. Miner FC. If two heads are better than one, why do I have bruises on my forehead? Managing the group process. *Clinical laboratory management review : official publication of the Clinical Laboratory Management Association* 1991;5(5):386-93.

134. Perry JM. Multidisciplinary shared leadership. *Journal for Healthcare Quality: Promoting Excellence in Healthcare* 2000;22(3):18-21.

135. Randal J. Are ethics committees alive and well? *The Hastings Center report* 1983;13(6):10-12.

136. Seekins T, Mathews RM, Fawcett SB. Enhancing leadership skills for community self-help organizations through behavioral instruction. *Journal of community psychology* 1984;12(2):155-63.

137. Shampain P. Committee dynamics. Part II: Taking command of the medical staff committee process. *QRC advisor* 1989;5(10):1-5.

138. Watson REL. The Role of the Department Chair. 1986

139. Whitman NI. The committee meeting alternative: Using the Delphi technique. *Journal of Nursing Administration* 1990;20(7-8):30-36.

Study design

140. Alvarez M, Kolehmainen CJ, Baier JM, Roman-Rosado G, Tippie R, Uptegraw JE, O'Hearn H, Serwe-Behnke A, Vogelman B, Carnes M, Hol-Land R. Choose your own adventure in interprofessional education. *Journal of General Internal Medicine* 2019;34(2 Supplement):S793.

141. Anderson S. Using Aboriginal governance and engagement to support cancer research. *Asia-Pacific Journal of Clinical Oncology* 2021;17(SUPPL 5):25.

142. Armstrong PW, Califf RM. Data and safety monitoring boards: Academic credit where credit Is due? *JAMA: Journal of the American Medical Association* 2013;310(15):1563-64.

143. Armstrong SM, Sanford K. Developing leaders: Succession planning involves everyone. *Transfusion* 2010;50(SUPPL. 2):243A-44A.

144. Auger J, Lee K, Haley R, Scott V. Core facility management models: Development and culture. *Journal of Biomolecular Techniques* 2009;20(1):24.

145. Bains W. Leadership and innovation. How consensus management blocks genuine innovation. *Bioscience Hypotheses* 2009;2(5):277-81.

146. Barza E. Nurses in leadership and decision-making roles: New I.R. Model. *Journal of Radiology Nursing* 2012;31(2):69.

147. Belle-Isle L, Pauly B, Benoit C. At the table with people who use drugs: How is power in decision-making being shared? *Canadian Journal of Infectious Diseases and Medical Microbiology* 2015;26(SUPPL. SB):119B.

148. Berends L. Ethical decision-making in evaluation. *Evaluation Journal of Australasia* 2007;7(2):40-45.

149. Bisol S, Roy A. Leveraging shared leadership in the sterile processing department to engage staff in process improvement. *American Journal of Infection Control* 2014;42(6 SUPPL. 1):S69-S70.

150. Cambor C, Roth DB, Loundas C, Heuer M, Watt CD, Montone K, Sesok-Pizzini D. Development of a dynamic leadership curriculum to enhance residency education in pathology and laboratory medicine. *Laboratory Investigation* 2016;96(SUPPL. 1):140A.

151. Colones R. Team effort. McLeod uses 'patient rounds,' leadership meetings to boost quality. *Modern healthcare* 2005;35(6):24-25.

152. Csaszar F, Enrione A. When Consensus Hurts the Company. *MIT Sloan Management Review* 2015;56(3):17-20.

153. Daykin A, Heawood A, Lane A, Macefield R, Gamble C, McCann S, Shorter G, Sydes MR. An ethnographic study of group decision making to understand and improve how trial steering committees contribute to trial conduct. *Trials* 2013;14(SUPPL. 1):82DUMMY.

154. Fiks AG, Kelly MK, Nwokeji U, Ramachandran J, Ray KN, Gozal D. A Pediatric Telemedicine Research Agenda: Another Important Task for Pediatric Chairs. *J Pediatr* 2022;251:40-43 e3.

155. Freeman M. Consensus is primary to group facilitation. *Group Facilitation* 2002 (4):56.

156. Girod SC. How to be an effective team leader and committee member or chair. *The academic medicine handbook: A guide to achievement and fulfillment for academic faculty* 2013 doi: 10.1007/978-1-4614-5693-3_38:309-14.

157. Harolds J. Planning and conducting meetings effectively, part I: Planning a meeting. *Clinical Nuclear Medicine* 2011;36(12):1106-08.

158. Harolds JA. Planning and conducting meetings effectively, Part II: Some component aspects of a meeting. *Clinical Nuclear Medicine* 2012;37(1):71-73.

159. Hartley J, Stansfield A. Leading through agonistic conflict: Contested sense-making in national political arenas. *Leadership* 2021;17(2):131-53.

160. Hash RB, Weintraut RJ, Gabriel SA, Treadwell TW, Fossum G. Developing departmental consensus in the search for a new chair. *Academic medicine : journal of the Association of American Medical Colleges* 2001;76(8):759-60.

161. Hauer KE, Cate OT, Boscardin CK, Iobst W, Holmboe ES, Chesluk B, Baron RB, O'Sullivan PS. Ensuring Resident Competence: A Narrative Review of the Literature on Group Decision Making to Inform the Work of Clinical Competency Committees. *Journal of graduate medical education* 2016;8(2):156-64.

162. Jaeger HF, Jaeger HF. Brigadier-General, Hilary F. Jaeger, MD, CHE, Chair of CCHSE's Ethics Committee, answers the question: How can a health service leader, such as a Regional Health Authority Chief Executive Officer, ensure that he or she remains compliant with the College's Code of Ethics when faced with political or fiscal pressure leading to management decisions which, in the opinion of the leader, are likely to adversely affect real health outcomes? *Healthcare Management Forum* 2008;21(3):44-45.

163. Kamau C, Harorimana D. Does knowledge sharing and withholding of information in organizational committees affect quality of group decision making? *Proceedings of the European Conference on Knowledge Management, ECKM* 2008:341-48.

164. Klinedinst R, Dingfield L, Farabelli JP, Ganta N. Balancing Democracy with Efficiency: Strategies for Leading an Interdisciplinary Palliative Care Team (SA505). *Journal of Pain and Symptom Management* 2020;59(2):493-94.

165. Kreamer L, Rogelberg S. Leadership & Professional Development: Evidence-Based Strategies to Make Team Meetings More Effective. *Journal of Hospital Medicine* 2020;15(4):236.

166. Li SA, Alexander P, Reljic T, Cuker A, Robby N, Wiercioch W, Guyatt G, Schunemann H, Djulbegovic B. The influence of activity roles and use of a structured framework on developing hematology clinical practice guidelines. *Blood* 2018;132(Suppl. 1)

167. Medvec VH, Berger G, Liljenquist K, Neale MA. Is a Meeting Worth the Time? Barriers to Effective Group Decision Making in Organizations. *Research on Managing Groups and Teams* 2003;6:213-33.

168. Nicholson D. Universal health coverage: Reaching a consensus. *The Lancet* 2015;385(9971):838.

169. Smales LA, Apergis N. The influence of FOMC member characteristics on the monetary policy decision-making process. *Journal of Banking and Finance* 2016;64:216-31.

170. Taylor C, Shewbridge A, Harris J, Green JS. Benefits of multidisciplinary teamwork in the management of breast cancer. *Breast Cancer: Targets and Therapy* 2013;5:79-85.

Government/legal/policy debate

171. Claussen CA, Matsen E, Røisland Ø, Torvik R. Overconfidence, monetary policy committees and chairman dominance. *Journal of Economic Behavior & Organization* 2012;81(2):699-711.

172. Meade EE. The FOMC: Preferences, voting, and consensus. *Federal Reserve Bank of St Louis Review* 2005;87(2 I):93-101.

173. Peggy S. EDUCATION GOVERNANCE REFORM IN ONTARIO: NEOLIBERALISM IN CONTEXT1. 2012

174. Riboni A, Ruge-Murcia FJ. Monetary policy by committee: Consensus, chairman dominance, or simple majority? *Quarterly Journal of Economics* 2010;125(1):363-416.

175. Russell A, Wainwright M, Tilson M. Means and ENDS-e-cigarettes, the Framework Convention on Tobacco Control, and global health diplomacy in action. *Global Public Health* 2018;13(1):83-98.

176. Waring S, Moran JL, Page R. Decision-making in multiagency multiteam systems operating in extreme environments. *Journal of Occupational and Organizational Psychology* 2020;93(3):629-53.

No full text available

177. Adigun JO. Perceptions of the roles and responsibilities of it administrators regarding technology strategic plans for client support centers in Tennessee Board of Regents institutions. 2013;73

178. Injety R, Jones S, Pandian J, Sylaja P, Padma MV, Sharma S, Webster J, Kulkarni GB, Sharma A, Lightbody C, Watkins C, Georgiou R. IMPROVING SYSTEMS OF ETHICAL APPROVALS FOR MULTICENTRE STROKE STUDIES IN INDIA - INSIGHTS FROM THE INDIAN STROKE CLINICAL TRIAL NETWORK (INSTRUCT NETWORK). *International Journal of Stroke* 2023;18(3 Supplement):110-11.

179. Katz N. Visualizing research performance: bringing strategic insight to research management. *Library Connect Newsletter* 2009;7(1):7-7.

180. Mindy RW. The Board Chair Handbook.

Not published in English

181. Kumsura K, Yamnill S. Factors affecting the decision-making ability of the basic educational institution committee in secondary schools. *Kasetsart Journal - Social Sciences* 2011;32(1):66-78.
